# Evolutionary and phenotypic characterization of spike mutations in a new SARS-CoV-2 Lineage reveals two Variants of Interest

**DOI:** 10.1101/2021.03.08.21253075

**Authors:** Paula Ruiz-Rodriguez, Clara Francés-Gómez, Álvaro Chiner-Oms, Mariana G. López, Santiago Jiménez-Serrano, Irving Cancino-Muñoz, Paula Ruiz-Hueso, Manuela Torres-Puente, Maria Alma Bracho, Giuseppe D’Auria, Llúcia Martinez-Priego, Manuel Guerreiro, Marta Montero-Alonso, María Dolores Gómez, José Luis Piñana, SeqCOVID-SPAIN consortium, Fernando González-Candelas, Iñaki Comas, Alberto Marina, Ron Geller, Mireia Coscolla

## Abstract

Molecular epidemiology of SARS-CoV-2 aims to monitor the appearance of new variants with the potential to change the virulence or transmissibility of the virus. During the first year of SARS-CoV-2 evolution, numerous variants with possible public health impact have emerged. We have detected two mutations in the Spike protein at amino acid positions 1163 and 1167 that have appeared independently multiple times in different genetic backgrounds, indicating they may increase viral fitness. Interestingly, the majority of these sequences appear in transmission clusters, with the genotype encoding mutations at both positions increasing in frequency more than single-site mutants. This genetic outcome that we denote as Lineage B.1.177.637, belongs to clade 20E and includes 12 additional single nucleotide polymorphisms but no deletions with respect to the reference genome (first sequence in Wuhan). B.1.177.637 appeared after the first wave of the epidemic in Spain, and subsequently spread to eight additional countries, increasing in frequency among sequences in public databases. Positions 1163 and 1167 of the Spike protein are situated in the HR2 domain, which is implicated in the fusion of the host and viral membranes. To better understand the effect of these mutations on the virus, we examined whether B.1.177.637 altered infectivity, thermal stability, or antibody sensitivity. Unexpectedly, we observed reduced infectivity of this variant relative to the ancestral 20E variant *in vitro* while the levels of viral RNA in nasopharyngeal swabs did not vary significantly. In addition, we found the mutations do not impact thermal stability or antibody susceptibility in vaccinated individuals but display a moderate reduction in sensitivity to neutralization by convalescent sera from early stages of the pandemic. Altogether, this lineage could be considered a Variant of Interest (VOI), we denote VOI1163.7. Finally, we detected a sub-cluster of sequences within VOI1163.7 that have acquired two additional changes previously associated with antibody escape and it could be identified as VOI1163.7.V2. Overall, we have detected the spread of a new Spike variant that may be advantageous to the virus and whose continuous transmission poses risks by the acquisition of additional mutations that could affect pre-existing immunity.

## Introduction

Genomic surveillance of viral mutations is the first step in detecting viral changes that could impact public health by interfering with diagnostics, modifying pathogenicity, or altering susceptibility to existing immunity or treatments. In many countries, the challenge of detecting new mutations of interest in SARS-CoV-2 is approached by sequencing representative genomes from circulating viruses, sharing sequence information on public databases (e.g. GISAID^1^), and analysing them in real-time using platforms, such as Nextstrain^2^. While mutations appear randomly, their fate in the population depends on a combination of the conferred fitness advantage as well as stochastic and demographic processes. A first step in assessing the potential public health impact of mutations is to decipher if their increase in frequency is due to chance or adaptation. If found to be adaptive, it is important to evaluate whether their adaptation is linked to an improved ability to replicate, colonize, transmit, or evade antiviral hosts defences^3^. An important challenge in the field is to decipher which of all the variants that appear should be monitored to implement measures to mitigate their risk to public health. Genotypes that are phenotypically different from a reference isolate or have mutations that lead to changes associated with either established or suspected phenotypes could be considered Variants of Interest (VOI) if they also fit one of the following criteria: i) cause community transmission/multiple COVID-19 clusters or ii) have been detected in multiple countries^4^. Among VOI, only those genotypes that are associated with higher transmissibility, with detrimental changes in COVID-19 epidemiology, with increased virulence, with changes to clinical presentation, or with decrease effectiveness of public health measures, diagnostics, vaccines or therapeutics are further categorized as variants of concern^4^.

Mutations in SARS-CoV-2 have been reported since the early stages of the epidemic^5–7^. While no signs of recombination have been detected so far among SARS-CoV-2 variants^8^, the most common mutations described are single nucleotide polymorphisms (SNPs) and small deletions^9–11^. Genomic surveillance of mutations has been mostly focused on the Spike (S) protein because of its key roles in viral entry and immunity^12^, as well as the fact that this protein constitutes the basis of numerous SARS-CoV-2 vaccines^13^. S is a homotrimeric protein, whose heavily glycosylated ectodomain protrudes from the viral membrane, showing a bat-like shape with a N-terminal globular head portion connected to the membrane by an elongated stalk^14^. The S protein is proteolytically processed by the cellular furin protease into the S1 and S2 subunits^15, 16^. Additional proteolytic cleavage occurs following S protein binding to the host receptors, facilitating S1 subunit release. The C-terminal S2 subunit remains trimeric in the viral membrane but undergoes conformational changes that promote viral membrane fusion with the host cell^17^. A key role in these conformational changes is played by two heptad repeat motifs, HR1 and HR2 that, starting from the head and stalk regions in the pre-fusion state of S protein, form a HR1-HR2 six-helix bundle in the post-fusion state that is critical for viral entry^18^.

The first mutation that was identified as of potential concern was an aspartic acid to a glycine mutation in the S1 subunit of the S protein at position 614 (D614G). D614G emerged early in the epidemic, became predominant in most countries within 2 months, and completely dominated the epidemic by August 2020^19^. As with any mutant, the initial spread of this mutation could have resulted from stochastic events, the dynamics of epidemic, or an intrinsically higher viral fitness. More than six months after the initial report of this mutation, several studies have found evidence in favour of higher transmission efficacy in animal models and human populations^5, 20–22^. This variant replicates better in some cell culture and animal models^20, 21, 23^, and is associated with higher viral loads in infected individuals^19^; importantly, however, it does not impact diagnostics or vaccine efficacy.

Following the first wave of the pandemic, additional variants have been reported from many countries. Among the first of these was the A222V mutation at the N-terminal domain (NTD) of the S1 subunit, which occurred in the background of the D614G S protein mutation. This variant, termed 20E, was first sequenced in Spain and expanded throughout Europe^6^. Other variants have been reported since, such as the so-called “cluster 5”, which harbours a combination of 3 SNPs and single deletion related to mink farms in Denmark^24^. One of the SNPs is in the S protein of this variant, Y453F, occurs in the receptor binding domain (RBD) and may increase binding to cell receptors in mink^25^. Transmission of this variant between humans and minks has been reported^7^, highlighting a possible risk of expansion in the human population, which resulted in proposals for large scale culling of mink populations in Denmark. Studies to assess the biological impact of this mutant have not been reported but there is no evidence for its wide spread over the course of >6 months since its description^26^. By December 2020, three variants of concern (VOCs) were described, all of which share the N501Y amino acid replacement in the RBD of the S protein: 20I/501Y.V1 (also called Lineage B.1.1.7) was originally described in the UK^11^, 20H/501Y.V2 (B.1.351) in South Africa, and 20J/501Y.V3 (P1) in Brazil. These variants are of particular concern because they are more transmissible^27–29^ and, although data on antigenicity and disease severity is not conclusive, could result in reduced susceptibility to neutralization by existing immunity and affect vaccine efficacy^30–33^. Importantly, all of these variants have spread outside of the country where they were initially identified and are estimated to spread faster than other co-circulating genotypes^27, 34, 35^.

In this work, we have performed a detailed phylogenomic analysis of the appearance, spread and evolution of two mutations involving amino acid positions 1163 and 1167 of the HR2 functional motif of S protein. Our results provide evidence of repeated, independent emergence, suggesting these mutations contribute to increased viral fitness. In addition, we have evaluated the biological relevance of these mutations to viral infectivity, virion stability, and neutralization by sera from convalescent and vaccinated individuals.

## Results

### Multiple and independent mutations in amino acid positions 1163 and 1167 of the Spike protein

SARS-CoV-2 genetic variation has been monitored by the Spanish sequencing consortium SeqCOVID to follow the expansion of mutations that could potentially result in a change of the biological properties of the virus. We focused on mutations in the S protein because of its relevance for infection and immunity^12^. We detected two mutations in the *spike* gene: G25049T (D1163Y) and G25062T (G1167V), which appeared in Spain as early as March and April 2020, respectively (Supplementary Fig.1). These mutations continued arising independently of each other and, by the end of June, when the predominant circulating genotypes from the first wave in Spain had already been replaced by other variants^36^, were also observed together (Supplementary Fig.1). Both positions have mutated multiple times independently and to different amino acids at a lower frequency. On the one hand, D1163 appears mutated at least 99 times (D1163Y: 84, D1163V: 4, D1163G: 3, D1163A: 2, D1163E: 2, D1163H: 2, D1163N: 1, and D1163H/Y: 1) in 47 lineages according to the PANGO scheme^37^. On the other hand, G1167 appears mutated at least 54 times (G1167V: 39, G1167D: 4, G1167C: 3, G1167R: 3, G1167S: 3, G1167F: 1, and G1167A: 1) in 20 PANGO lineages including B.1 (Supplementary Fig.2e) and its derivatives B.26, B.40 (Supplementary Fig.2c), and D.2 (Supplementary Fig.2f).

**Figure 1.**
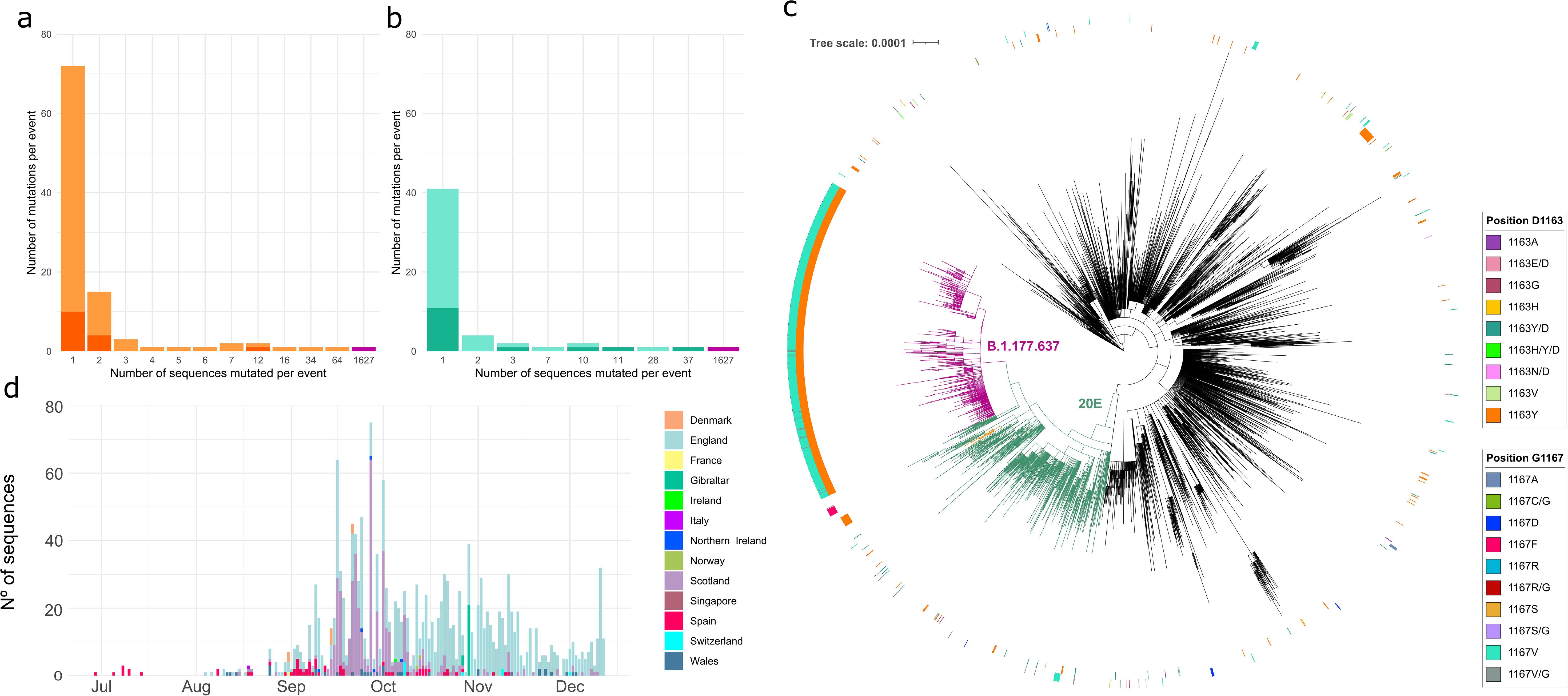
Sequences mutated at positions 1163 and 1167 of the S protein. **a.** The number of mutation events for amino acid replacement D1163Y (light orange) or another D1163 amino acid replacements (dark orange). **b**. The number of mutation events for amino acid replacement G1167V (light turquoise) or another G1167 amino acid replacements (dark turquoise). Bars coloured in magenta indicate the appearance of both the D1163Y and G1167V amino acid replacements in the same sequences. **c.** Maximum-likelihood phylogeny of 10,450 SARS-CoV-2 genomes. The inner circle of the rings represents sequences with amino acid changes in position D1163 of the S protein. The external circle represents sequences with amino acid changes in position G1167 of the S protein. Branches are coloured in magenta for B.1.177.637, green for clade 20E, and orange for cluster 1163.654. The scale bar indicates the number of nucleotide substitutions per site. **d.** Temporal distribution and frequency of sequences with variant B.1.177.637 coloured by geographical origin.

### Clusters of transmission with amino acid changes in positions 1163 and 1167 of Spike

Positions 1163 and 1167 in the S protein have mutated independently multiple times in SARS-CoV-2. The majority of mutated sequences (94.43%) were found in transmission clusters (see methods for definition of clusters; Figure 1a,b), with a small minority not belonging to a transmission cluster due to either incomplete sampling or failure to spread. While different amino acids changes have been detected at both positions, only one change at each position appeared in most clusters: D1163Y in 83.33% and G1167V in 69.23% clusters. D1163Y appeared in 22 transmission clusters (Figure 1a) and G1167V in 8 clusters (Figure 1b). Interestingly, the biggest cluster included both the D1163Y and G1167V mutations together. We denote this cluster as B.1.177.637, which was detected in 65 sequences from Spain until December 2020, representing 1.17% of the Spanish sequences. Globally, this cluster, which is within lineage 20E (also described as 20A.EU1^6^, and B.1.177^37^), includes 1,627 sequences (Figure 1c). B.1.177.637 is characterized by nine nonsynonymous and six synonymous mutations with respect to the reference sequence from Wuhan (Supplementary table 2), but no deletions were shared among B.1.177.637 sequences. Amino acid changes were found in A222V, D614G, D1163Y, and G1167V in the S protein, A220V and P365S in the N protein, V30L in ORF10, L67F in ORF14, and P4715L in ORF1ab (Supplementary Fig.3 and Supplementary table 2). Synonymous mutations were also observed in the *orf1ab*, *n* and *m* genes (Supplementary Fig.3 and Supplementary table 2). All evidence support we consider this cluster a new Lineage, and we requested to be under the name B.1.177.637 (https://github.com/cov-lineages/pango-designation/issues/22).

**Figure 3.**
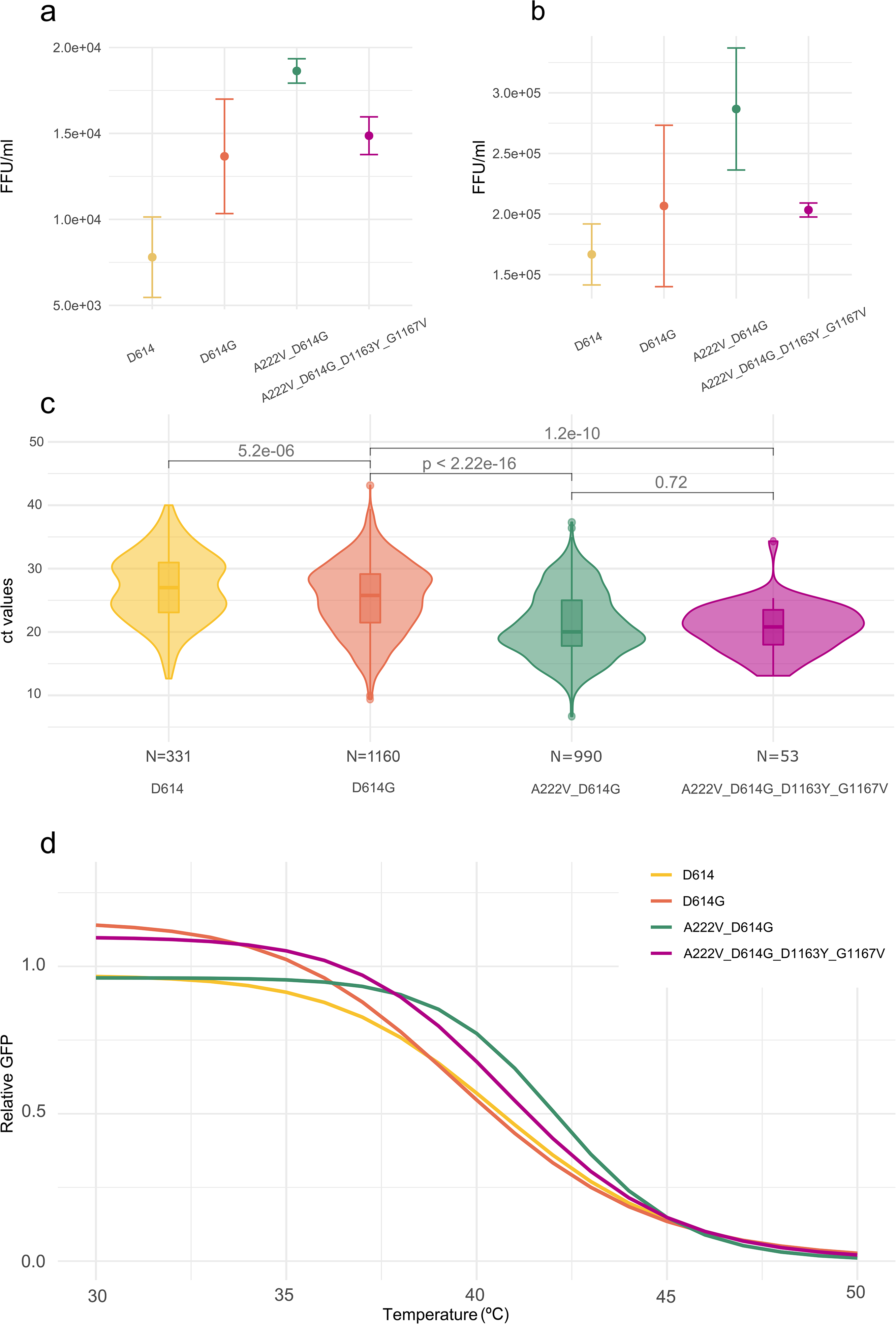
Comparison of the infectivity and stability of different S genotypes. **a, b.** The infectivity of VSV particles pseudotyped with each S protein genotype in either Vero (**a**) or human A549 cells expressing ACE2 and TMPRSS2 (**b**). The mean and standard deviation of three replicates is plotted. **c.** Comparison of cycle threshold (Ct) values for the *n* gene from patients infected with viruses encoding different S protein variants. Data is derived from 2,534 sequences from SeqCOVID consortium. The number of observations (N) analysed for each genotype is indicated. **d.** The thermal sensitivity of VSV pseudotyped with different S genotypes following incubation at 15 minutes. Data are standardized to the surviving fraction following incubation at 30°C, and the three-parameter log-logistic equation is plotted. FFU: focus forming units.

Within 20E, the second largest cluster including any of these mutations was observed in 34 sequences with E654Q and D1163Y in S protein plus 7 nonsynonymous and 6 synonymous mutations (Supplementary table 2 and Supplementary Fig.3). We denote this second cluster, which is also embedded within lineage 20E, cluster 1163.654 (Figures 1c and S3). Cluster 1163.654 appeared first in Ireland on 2020-07-23 and subsequently appeared in Spain and England. However, after three months, cluster 1163.654 is no longer being detected. A large cluster within Lineage 20E is formed by 37 sequences with the mutation G1167F. Sequences for this cluster were obtained in Wales between the end of October and the beginning of November 2020 (dark pink in the external circle in Supplementary Fig.2g).

We found additional clusters involving mutations in position 1163 of the S protein; the largest cluster is composed of 64 sequences within Lineage B.1. The majority of which are from Denmark but also includes sequences from England and Sweden (orange in external ring in Supplementary Fig.2e). Among other smaller clusters, a cluster of 10 sequences with G1167V (within Lineage B.1) appeared in Spain in March (indicated in cyan within the external circle of Supplementary Fig.2e). The cluster was detected in two regions in Spain (Valencia, and Galicia), but was controlled with lockdown measures imposed in Spain from March to May and was not detected after May 2020. The same mutation was found in a cluster of 28 sequences within 20E from England and Wales between October and November 2020 (indicated in cyan within the external circle of Supplementary Fig.2g). Similarly, another cluster of 11 sequences in Lineage B.1.1 and with the mutation G1167A was detected in early January 2021, encompassing sequences from Ecuador, Colombia, and Peru (Supplementary Fig.2f). Within Lineage B.53, we found a cluster of 12 sequences from Lithuania with mutation D1163Y. Finally, we found small clusters with mutations in 1163 or 1167 within Lineage B.40 and Lineage A (Supplementary Fig.2b, c).

Because of the risk posed by VOC^11, 26, 35^, we examined whether mutations in 1163 and 1167 of the S protein were observed in the three VOC described to date. Indeed, mutations in these two positions were observed in two VOC, 20I/501Y.V1 and 20H/501Y.V2. Mutations involving amino acid positions 1163 and 1167 appeared independently in the background of 20I/501Y.V1 multiple times. Specifically, mutations in D1163 have occurred at least 13 times in 24 sequences, including four transmission clusters and nine unique sequences (Supplementary Fig.4), while mutations at G1167 were observed in at least five independent sequences. Interestingly, D1163Y and G1167V were observed together in only one individual within 20I/501Y.V1, although they were not fixed (relative frequency of 27% and 17% of the reads with D1163Y and G1167V, respectively; Supplementary table 3). Finally, only two sequences that harbour the amino acid replacement G1167V in the S protein were observed in the genomic background 20H/501Y.V2 (Supplementary table 3).

### Evolution of B.1.177.637

We explored the emergence and evolution of B.1.177.637, the largest and most successful cluster involving amino acid changes in positions 1163 and 1167 of the S protein. Lineage B.1.177.637 appeared in Spain in June 2020 in sequences from the Basque Country (Figure 1d, and Supplementary Video 1) and subsequently appeared in individuals from other countries, comprising a total of 1,627 sequences in GISAID (0.60% of 270,869 analysed sequences by 23^rd^ of December 2020) (Figure 1d and Supplementary Video 1). The majority of the B.1.177.637 sequences were obtained from the United Kingdom, including England (1,058), Scotland (419), Wales (34) and Northern Ireland (5), but were also observed in Gibraltar (24 sequences), indicating successful migration and transmission (Supplementary Video 1). Although B.1.177.637 is not well represented in sequences from other countries, it has been found in multiple sequences from Denmark (9), Switzerland (8), Norway (2), and single sequences from Italy, France, Singapore, and Ireland. By the end of 2020, B.1.177.637 was still circulating in Europe (Figure 1d and Supplementary Video 1), and by the end of February 2021 it was represented by 1,923 sequences in GISAID (0.33% of submitted sequences).

Within B.1.177.637, we detected additional SNPs in individual sequences or small groups of sequences. One of these changes is E484K in the RDB of the S protein, a mutation present in three VOCs (20I/501Y.V1, 20H/501Y.V2 and 20J/501Y.V3) that is implicated in increased ACE2 binding^38^ and reduced neutralization by antibodies^39^. In addition, we found another change associated with evasion of antibody immunity: a deletion of positions 141-144 in the S protein, which partially overlaps with a smaller deletion at 144 reported in VOC 20I/501Y.V1^40^. This sub-cluster included five sequences along January 2021 from England and Wales (Supplementary table 2). The five sequences formed a monophyletic group embedded in B.1.177.637 (Supplementary Fig.5), identified as cluster B.1.177.637.V2, which displays other nonsynonymous and synonymous mutations (Supplementary table 2) and only two sites are polymorphic within B.1.177.637.V2.

**Figure 4.**
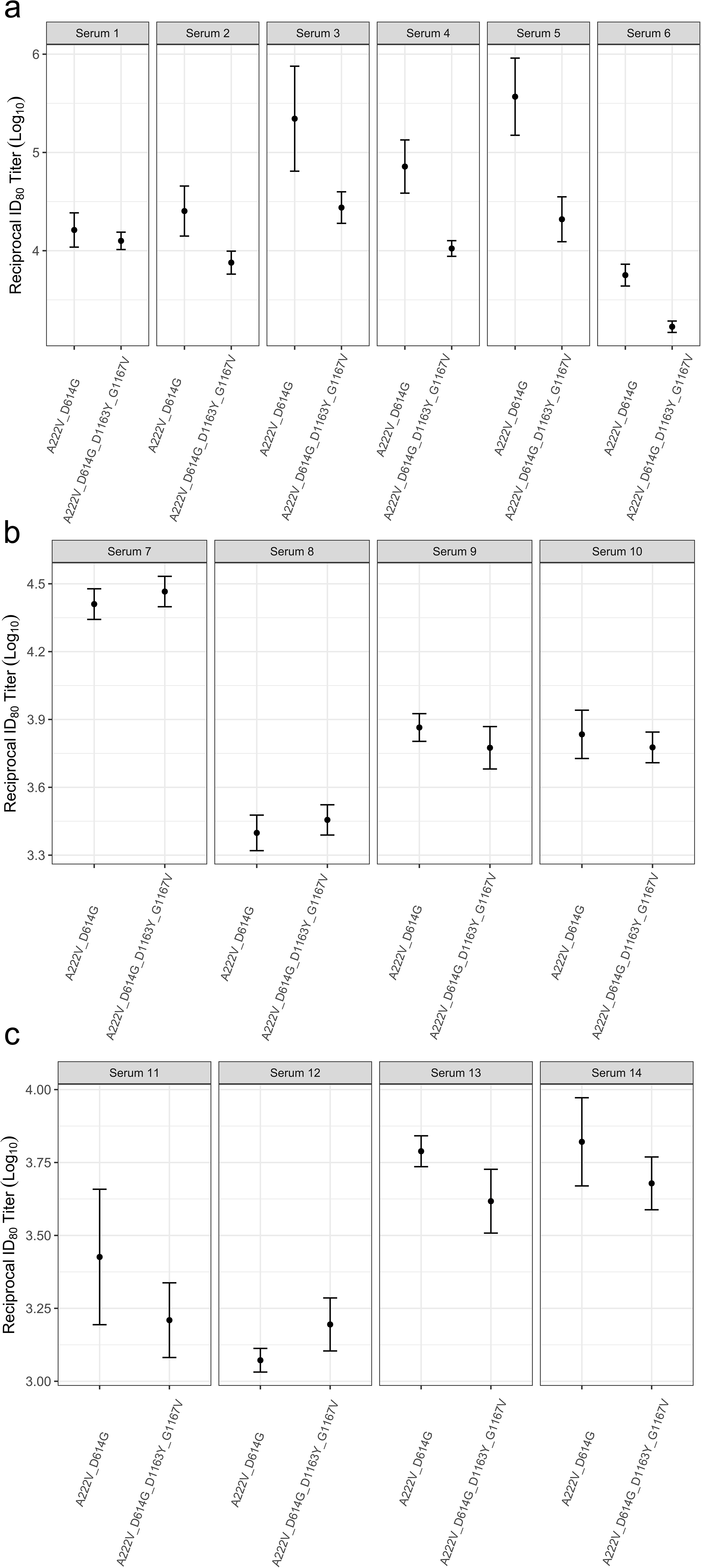
Antibody neutralization of 20E and B.1.177.637 variants. The reciprocal titer at which infection with the 20E S genotype (A222V and D614G) or B.1.177.637 S genotype (20E plus D1163Y and G1167V) is reduced by 80% (ID80) by sera from individuals infected during the early stage of the pandemic in (**a**) or during a later stage of the pandemic (**b**) or from donors vaccinated with the BNT162b2 vaccine (**c**). The mean and standard error of three replicates is plotted.

### Positions 1163 and 1167 of the S protein are located in the heptad repeat 2 motif

The S protein mediates both the binding to cellular receptors and entry into the host cells^12^. For the former, the RBD motif in the S1 subunit interacts with the cellular receptor in the pre-fusion state. In the post-fusion state, two heptads repeat sequences (HR1 and HR2) in the S2 subunit must form a six-helix bundle in order to bring the viral and cellular membrane into close proximity^41, 42^ (Figure 2). S protein positions 1163 and 1167 are both located within the HR2 domain. Specifically, 1167 is present at the beginning of the HR2 motif and 1163 in its upstream linker region (Figure 2a). Interestingly, this motif is highly invariable, showing 100% conservation across 14 related sarbecoviruses to which SARS-CoV-2 belongs to (Supplementary table 1)^43, 44^. Structural characterization of full-length ectodomain of S protein has shown that the stalk portion encompassing positions 1163 and 1167 presents intrinsic flexibility in the pre-fusion state^18^, precluding its atomic visualization. This has been recently confirmed by high-resolution cryo-electron tomographic reconstitution of SARS-CoV-2^14^, where this region was observed to constitute a flexible hinge that acts as a “knee”, connecting two helical coiled-coil regions of the stalk (upper and lower legs; Figure 2b). Within this structure, the conformational freedom provided by the glycine residue at position 1163 should play a key role in the flexibility of the knee. In contrast, in the post-fusion state, this region shows high rigidity due to a strong structural rearrangement of the HR2 motif, which adopts an extended conformation and tightly packs along the central 3-helix bundle stem formed by the HR1 motif (Figure 2c). The resulting HR1-HR2 bundle plays a key role in the mechanism of viral-host membrane fusion^18, 45^ and mutations in this region could have significant impact on the function of the S protein. In addition, the HR2 region is highly glycosylated, which is observed to be regularly spaced in both the pre- and post-fusion states and to mostly align to the side of the helix bundle^14, 18, 45^. Of note, two of these branched sugars are placed at positions N1158 and N1173, hiding positions 1163 and 1167 (Figure 2b). Therefore, changes in stalk flexibility might have relevance in immunity by influencing both the intrinsic degree of exposure of this region and its sugar shielding.

**Figure 2.**
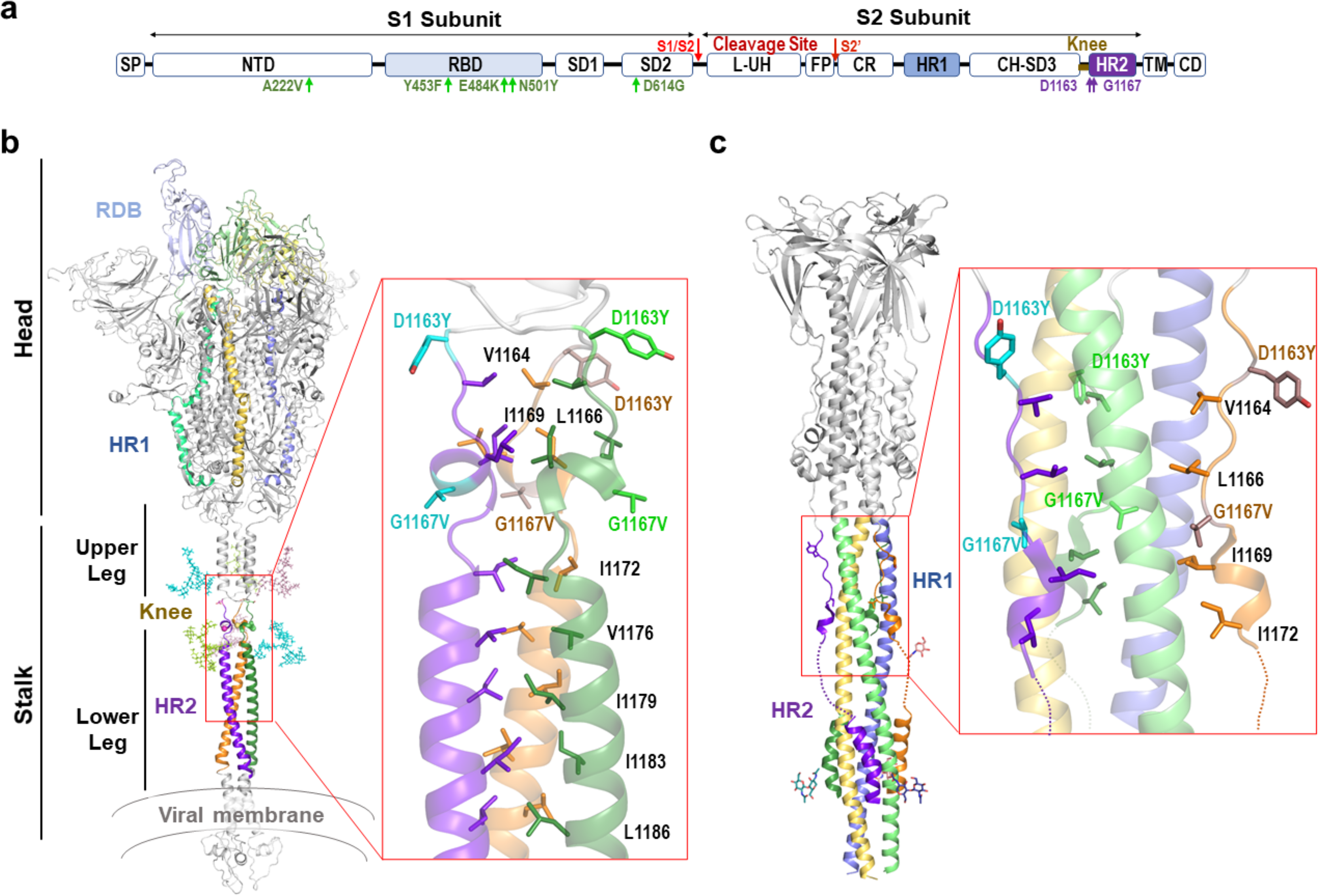
The structure of 1163 and 1167 in the pre and post-fusion states of S protein. **a.** Schematic representation of the S protein. SP. Signal peptide; NTD, N-terminal domain; RBD, receptor-binding domain; SD1-2, subdomains 1 and 2; L-UH, Linker-Upstream helix; FP, fusion peptide; CR, connecting region; HR1, heptad repeat 1; CH-SD3, central helix subdomain 3; BH, β-hairpin; HR2, heptad repeat 2; TM, transmembrane; CD, cytoplasmic domain. Mutations D1163Y and G1167V are indicated in purple and other mutations described in the text in green. **b.** Cartoon representation (*left*) of a structural model of pre-fusion membrane-bound trimeric S protein^46^. In each subunit the RBD, HR1 and HR2 domains are coloured in different tones (light to dark) of blue, yellow, and green. The N-glycosylation of N1155 and N1176 are shown in stick representation and coloured as the corresponding subunit. Functional and structural regions are marked. A close-view (*right*) of the N-terminal portion of HR2 where D1163Y and G1167V mutations are found. The side-chains of mutated and hydrophobic residues in the HR2 region are shown in stick representation and coloured as corresponding subunit (mutated residues in lighter tone). **c.** Cartoon representation (left) of S2 subunit in post-fusion conformation with HR1 and HR2 regions coloured as in b and N-glycosylation around mutation position shown as sticks. A close-view of the region encompassing the mutations (right), showing in stick representation the mutated and hydrophobic residues from the HR2 region shown in panel b. Dotted lines highlight HR2 disordered regions in the Cryo-EM structure.

Using the available structural information of the S protein in the pre- and post-fusion conformations^18^, we examined the possible implications of these mutations to viral infectivity. Based on these structures, G1167V mutation is predicted to confer significant rigidity to the structure in two ways. First, the introduction of a side chain strongly reduces the conformational freedom provided by the glycine residue. Second, the presence of the new aliphatic side chain provided by the valine residue strongly increases hydrophobicity, likely promoting the burial of this side chain in the HR1 helix 3-bundle stem in the post-fusion state or favouring its integration in the neighbour helical coiled-coil in the pre-fusion state (Figure 2b,c). Unlike position 1163, position 1167 is fully exposed to the solvent in both the pre- and post-fusion states (Figure 2b,c). Hence, the effect of D1163Y is likely to stem a change in nature of the side chain, switching from a charged aspartic acid residue at physiological pH to a polar group with hydrophobic properties in the tyrosine.

### Spike aminoacid changes D1163Y and G1167V do not increase viral infectivity

Previous reports have indicated that mutations in the S protein can increase infectivity^5, 21, 47–49^. Because the highest transmission success for mutations in S positions 1163 and 1167 corresponds to the double mutant D1163Y and G1167V (characteristic of B.1.177.637), we explored whether these mutations in combination have an influence on infectivity. For this, we pseudotyped vesicular stomatitis virus lacking its glycoprotein and encoding GFP^50^ (VSVΔG-GFP) with different S genotypes: Wuhan (D614), D614G, 20E (A222V and D614G), or B.1.177.637 (A222V, D614G, D1163Y and G1167V). Infectious virus production was then assessed by limiting dilution and counting of GFP-positive cells in both Vero cells and A549 human alveolar basal epithelial cells expressing human ACE2 and TMPRSS2 (A549-hACE2-TMPRSS2). As previously reported^19, 21, 51^, the 20E S genotype enhanced infectivity relative to the Wuhan S genotype by 70% in both Vero (p-value = 0.005 by unpaired t-test; Figure 3a) and A549-hACE2-TMPRSS2 cells (p-value = 0.016 by unpaired t-test; Figure 3b). The 20E S genotype also showed a trend towards increased infectivity versus the D614G mutation alone (35% increase in both cell lines), as has been previously reported^49^, yet the difference was not statistically significant (p-value > 0.05 by unpaired t-test; Figure 3a,b). In contrast, B.1.177.637 S genotype significantly diminished virus infectivity versus the 20E genotype, reducing virus titers by 20% in Vero cells (p-value = 0.009 by unpaired t-test; Figure 3a) and 29% in A549-hACE2-TMPRSS2 (p-value = 0.03 by unpaired t-test; Figure 3b). This is in agreement with a potential stabilization of the HR2 helix (Figure 2), which should limit the ability of the S protein to sample different structural conformations that could be required for binding host-cell receptors. Hence, B.1.177.637 S genotype does not increase infectivity *in vitro*.

To examine whether reduced infectivity could also be observed *in vivo*, we tested if individuals infected with B.1.177.637 have different viral loads. For this, we used the cycle threshold (Ct) of real-time PCR used for diagnosis as a surrogate. As previously reported^19^, we detected higher Ct values for D614 wild-type variant (Ct mean = 27.00) compared to genotypes encoding the D614G S protein mutation (Ct mean = 25.32; p-value < 0.01 by unpaired Wilcoxon test, Figure 3c). However, we did not find significant differences in viral loads between individuals infected with B.1.177.637 genotype and other genotypes within 20E (Ct mean = 21.14 vs Ct mean = 20.63, p-value = 0.72 by unpaired Wilcoxon test, Figure 3c). Interestingly, higher viral loads were observed in individuals infected with B.1.177.637 and others 20E (D614G and A222V) compared to the D614G alone (D614G Ct mean = 25.32, 20E Ct mean = 21.14, B.1.177.637 Ct mean = 20.63, p-value < 0.01 for both comparisons by unpaired Wilcoxon test, Figure 3c). This data suggests that, unlike the results of the in vitro studies, B.1.177.637 replicates as efficiently as other 20E genotypes *in vivo*, although mutations outside of the S protein could contribute to this result.

### Amino acid changes D1163Y and G1167V do not alter S protein stability

As increased S protein stability could impact transmissibility by maintaining virion infectivity during the intra-host transmission period, we assessed the temperature sensitivity of the different S variants. For this, we subjected VSV particles pseudotyped with different S genotypes to a range of temperatures for 15 minutes, after which we evaluated the surviving fraction. Overall, no major differences in the degree to which the different S proteins lost infectivity upon heat exposure were observed, with all S proteins showing a 50% reduction in infectivity at a similar temperature range (39.8-42.2°C; p-value > 0.05 for all except Wuhan S genotype (D614) versus 20E S genotype (A222V and D614G), where p-value = 0.01; Figure 3d). Hence, the D1163Y and G1167V mutations do not seem to have a major impact in the thermal stability of the S protein.

### Spike D1163Y and G1167V modestly reduce sensitivity to neutralization by existing antibody immunity

Positions 1163 and 1167 of the S protein have been reported to occur in both T and B cell SARS-CoV-2 epitopes^52–54^. Moreover, numerous studies have shown that mutations in the S protein can affect antibody neutralization^30, 31^. We therefore examined if the presence of D1163Y and G1167V alters the neutralization capacity of convalescent sera using VSV pseudotyped with either the 20E or B.1.177.637 S genotypes. In order to capture the potential influence of different infecting variants on antibody neutralization, we tested the sensitivity of these pseudotyped viruses to neutralization by sera from early (April 2020; First wave in Spain) or later (October 2020; Second wave in Spain) in the pandemic, when newer variants were dominant^6, 36^. Overall, B.1.177.637 genotype conferred a modest but statistically significant reduction in sensitivity to neutralization by six serum samples tested from the early stage of the pandemic, as measured by the titers required to inhibit viral entry by 80% (ID80; mean = 6.75, range: 1.30-17.68; p-value = 0.008 by paired t-test; Figure 4a). A statistically significant but smaller effect was observed when the titers required to inhibit viral entry by 50% were examined (ID50; mean = 2.27, range: 1.61-3.54; p-value < 0.001 by paired t-test; Supplementary Fig.6). In contrast, both 20E and B.1.177.637 were equally susceptible to sera from patients infected during the second wave (ID80; mean = 1.03, range: 0.87-1.23; p-value = 0.83 by paired t-test; Figure 4b). These results indicate that the D1163Y and G1167V mutations can provide some degree of escape from pre-existing antibody-based immunity relative to the 20E S genotype depending on the genomic background of the infecting genotype. As a modest reduction in titers was observed with sera from early in the pandemic (Figure 4a), which is more closely related to the current S genotype present in approved vaccines^55, 56^, we examined if B.1.177.637 S genotype resulted in reduced neutralization by sera from donors vaccinated with the BNT162b2 vaccine. No significant differences in susceptibility to antibody neutralization from vaccinated donors were observed between the two genotypes, indicating that VOI1163.7 is unlikely to alter the efficacy of vaccines based on the Wuhan S genotype (Figure 4c).

## Discussion

SARS-CoV-2 success is linked to its ability to infect and be transmitted. Mutations that emerge independently several times and increase in frequency are likely to confer enhanced viral infectivity, transmission, or immune evasion. The identification of such mutants is of great importance, as they can significantly impact public health. In this work, we have identified two mutations in the S protein that are likely to be beneficial for the virus based on several lines of evidence. First, these mutations are highly variable within SARS-CoV-2 but conserved across the closely related coronaviruses. Second, the vast majority of sequences harbouring these mutations appear in clusters (Figure 1a and 1b). Third, the largest cluster, and therefore the most successful in terms of transmission, includes both mutations together (Figure 1a and 1b). Additionally, both positions have been reported as positively selected multiple times throughout the SARS-CoV-2 phylogeny indicating a fitness advantage^57^. Although either mutation in isolation could be advantageous, their co-occurrence in a large cluster that has been sustained for more than six months across Europe is suggestive of increased fitness when both mutations are present together.

Positions 1163 and 1167 are found in the HR2 domain of the S protein, adjacent to the transmembrane domain. Examination of available structural data suggests that G1167V might alter the flexibility of the S protein stalk by both restricting the conformational freedom normally conferred by the glycine residue and by introducing a hydrophobic side chain that will favour burial in the HR2 coiled-coil leucine zipper of the pre-fusion state (Figure 2). This extensive flexibility of S prefusion stalk seems to be unique to the SARS-CoV-2 S protein and has not been reported for other class I fusion proteins^18^. The stalk flexibility has been suggested to increase avidity for the host receptors by allowing the engagement of multiple S proteins^18^. Therefore, stalk stabilization is likely to result in a reduced ability of S to bind receptors in the target cell. Indeed, we find the B.1.177.637 genotype to have reduced infectivity compared to the 20E genotype in both Vero and A549-hACE2-TMPRSS2 cells (Figure 3a, b). In contrast, viral load in individuals infected with different S genotypes indicated that 20E and B.1.177.637 reach similar degrees of viral replication in vivo (Figure 3c). This apparent discrepancy in the effect of the two mutations on infectivity may stem from the differences of SARS-CoV-2 infection *in vivo* and in the *in vitro* assay. A recent publication has suggested that the tyrosine-protein kinase receptor UFO (AXL) is an important mediator of SARS-CoV-2 entry and may be of higher relevance for infection of the lung than ACE2^58^. The effect of entry via AXL in the two cell lines used may be overshadowed by high levels of ACE2 expression. Alternatively, additional factors could underlie this difference, including the presence of additional mutations outside of the S protein or differences of viral loads across sampling times i*n vivo* during the infection.

Increased temperature stability can potentially confer a fitness advantage to the virus by reducing losses to infectivity during environmental transition between hosts. Hence, we also examined whether these mutations altered the temperature stability of the virions. Overall, no major difference in stability was observed between VSV pseudoparticles bearing the D614, D614G, 20E, or B.1.177.637 S protein genotypes (Figure 3d). Hence, changes in protein stability are unlikely to underlie the increased transmission of these variants.

Finally, as the S protein is a major target of the immune response^12^, immune evasion represents one possible consequence of mutations in this protein. Both S positions 1163 and 1167 are embedded in experimentally confirmed T cell and B cell epitopes. For T cell epitopes, a predicted HLA-II epitopes including position 1163 and 1167 has been experimentally verified to bind to HLA DRB1*01:01, the prototype molecule for the DR supertype (epitope identifier in Immune Epitope Data Base: 9006^59^). Additionally, D1163 is included in a SARS-CoV-2 T cell epitope eliciting T-cell responses in convalescent COVID-19 cases^60^ as well as in SARS-CoV-2-naïve individuals^53^, indicating cross-reactivity in epitopes involving these regions. B cell linear epitopes that span D1163 and G1167 have also been reported^52^, with D1163 belonging to a dominant linear B cell epitope recognized by more than 40% COVID-19 patients used in the assay^54^. D1163 is fully solvent exposed in available structure^18, 45^, making its side-chain easily accessible to antibodies, providing a potential mechanism for altering antibody binding. To directly examine whether the mutated S positions 1163 and 1167 influence susceptibility to pre-existing humoral immunity, we examined the neutralization capacity of convalescent sera against the 20E variant or B.1.177.637. For this, we used sera from both the first (April 2020) and second (October 2020) waves of the infection in Spain, because an almost complete replacement of SARS-CoV-2 S variants occurred between these two times of the pandemic in Spain^36^. When utilizing sera from donors infected during the first wave of the pandemic in Spain, we found a modest but statistically significant reduction in susceptibility to neutralization of the B.1.177.637 S genotype compared to the 20E S genotype of approximately 6-fold (Figure 4a). However, no difference in neutralization was observed between the two variants when sera from patients infected during the second wave was used (Figure 4b), highlighting variant-specific differences in antibody responses. Overall, the magnitude of the observed reduction in neutralization susceptibility to sera from individuals infected during the first wave was much less pronounced than that observed for other genotypes implicated in immune evasion^31^. Nevertheless, the degree of reduced neutralization required to confer a biologically relevant fitness advantage *in vivo* has not been established, and even relatively small reductions in susceptibility to antibody neutralizations could potentially confer a significant advantage to replication. Indeed, this has been suggested to be the case in an immunosuppressed individual treated with convalescent serum, where mutations selected during the course of treatment conferred a similar reduction to that observed in the current study^61^. Finally, no evidence was found for reduced neutralization by sera from donors immunized with the BNT162b2 vaccine (Figure 4c), which is based on the Wuhan S genotype, indicating antibody immunity elicited by Pfizer-BioNTech COVID-19 vaccine (BNT162b2; February 2021) would not be affected by amino acid replacements in 1163 and 1167 sites in S protein.

Although further experiments are needed to decipher the mechanisms by which the two S mutations identified could confer a selective advantage to the virus, the evidence presented supports B.1.177.637 as a VOI (VOI1163.7.V1) according to recently published criteria^4^. First, amino acid replacements in S protein: D1163Y and G1167V lead to changes associated with suspected phenotypic change because of the rigidity it poses to the S protein. Second, the genotype showed moderate but significantly lower antibody susceptibility compared to 20E S genotype. And third, it has been identified to cause community transmission, appearing in multiple COVID-19 cases, and detected in multiple countries. Additionally, a subgroup within B.1.177.637 (denoted as VOI1163.7.V2) includes two additional mutations leading to amino acid changes in S with established phenotypic impact on humoral immunity: E484K^62^ and 141-144Del^40, 63^.

Whether VOI1163.7.V1 and VOI1163.7.V2 will continue to increase in frequency and accumulate additional mutations that could improve its fitness and/or present challenges to vaccines or diagnostics remains to be seen. However, their characterization as VOI would help to discover if enough evidence holds to consider them VOC and therefore required monitoring.

## Methods

### Whole-genome sequencing and genome assembly of SeqCOVID consortium sequences

A total of 5,017 clinical samples were received, sequenced, and analysed by the SeqCOVID consortium from all autonomous communities of Spain. These samples were confirmed as SARS-CoV-2 positive by RT-PCR carried out by Clinical Microbiology Services from each hospital. Sequencing of the samples has been approved by the ethics committee: Comité Ético de Investigación de Salud Pública y Centro Superior de Investigación en Salud Pública (CEI DGSP-CSISP) N° 20200414/05. All sequences are available at GISAID under the accession numbers detailed in Supplementary table 4.

For sequencing, RNA samples were retro-transcribed into cDNA. SARS-CoV-2 complete genome amplification was performed in two multiplex PCR, according to the protocol developed by the ARTIC network^64^, using the V3 multiplex primers scheme^65^. From this step, two amplicon pools were prepared, combined, and used for library preparation. The genomic libraries were constructed with the Nextera DNA Flex Sample Preparation kit (Illumina Inc., San Diego, CA) according to the manufacturer’s protocol, with 5 cycles for indexing PCR. Whole-genome sequencing was performed in the MiSeq platform (2×200 cycles paired-end run; Illumina).

Reads obtained were processed through a bioinformatic pipeline based on iVar^66^, available at https://gitlab.com/fisabio-ngs/sars-cov2-mapping. The first step in the pipeline removed human reads with Kraken^67^; then fastq files were filtered using fastp^68^ v 0.20.1 (arguments employed: --cut tail, --cut-window-size, --cut-mean-quality, -max_len1, -max_len2). Finally, mapping and variant calling were performed with iVar v 1.2, and quality control assessment was carried out with MultiQC^69^.

### Analysis of the *spike* gene of sarbecoviruses related to SARS-CoV-2

14 sequences including SARS-COV-2 belonging to sarbecoviruses, sequences were annotated with annotation files available at NCBI database in order to locate the *spike* gene coordinates (accession numbers are available at supplemental table 1). For each sequence, nucleotide coordinates belonging to this gene were extracted with EMBOSS^70^. The 14 sequences harbouring the *spike* gene were concatenated and aligned with MEGA-X^71^ using amino acids with ClustalW algorithm with default options (alignment is available at https://github.com/PathoGenOmics/B.1.177.637_SARS-CoV-2)

### Sampling SARS-CoV-2 from non-Spanish consortium sequences

To build the global alignment, sequences were downloaded from GISAID^1^ including all the pandemic periods since the first known case sequenced (from 24 December 2019) until the last sample on 22 December 2020. We used two filters to select the dataset: sequences with more than 29,000 bp, and sequences with known dates of sampling. Sequences downloaded from GISAID were aligned against the SARS-CoV-2 reference genome^72^ using MAFFT^73^, omitting all insertions and getting an alignment length of 29,903 bp. The final alignment constructed included 270,869 sequences, all sequences with GISAID ID used for this study are available in Supplementary Table 5.

### Frequency and detection of mutated positions

Single nucleotide variants were detected using the global dataset alignment, generating a VCF file with SNP-sites^74^ v 2.5.1 (argument employed: -v), using the reference genome as the reference bases for detecting mutations. This VCF file was processed with a Python script (available at https://github.com/PathoGenOmics/B.1.177.637_SARS-CoV-2) to assess all mutated samples by position, calculating the frequencies of the global dataset and annotating sequences with the detected mutations. After that, the mutated positions were annotated with snpEff^75^ v 5.0 using SARS-CoV-2 reference (Wuhan first sequenced) database annotation (arguments employed: -c, -noStats, -no-downstream, -no-upstream, NC_045512.2).

Genotypes detected that involved mutations in 1163 and 1167 such as B.1.177.637 and cluster 163.654 were represented in a circos plot with the R package circlize^76^ v 0.4.12.1004. Nucleotide coordinates of SARS-CoV-2 are plotted in a non-closed circle, circle is annotated and coloured with genes of the virus, and mutated nucleotide positions of each genotype are connected between them through lines.

### Alignments

For the phylogenetic analysis, a reduced dataset was selected from the 270,869 sequences. Duplicated sequences were removed with seqkit v 0.13.2 (arguments employed: rmdup -s). 8,397 sequences were selected at random with the same temporal distribution by month as the initial dataset by Python scripting (available at https://github.com/PathoGenOmics/B.1.177.637_SARS-CoV-2). The 8,397 sequences were concatenated with 2,053 sequences selected as indicate above because harboured amino acid replacements in D1163 and G1167 of the S protein and resulted in an alignment of 10,450 sequences (Supplementary Table 6).

The dataset to represent 20I/501Y.V1 phylogenetic relationships include 3,067 randomly selected samples identified by Pangolin typing system (https://github.com/cov-lineages/pangolin) as lineage B.1.1.7 plus the 33 sequences with amino acid replacements in D1163 and/or G1167 (Supplementary Table 7).

For all the alignments, problematic positions reported by Lanfear, R.^77^ were masked for the phylogenetic reconstruction using masked_alignment.sh script.

### Phylogenetic analysis

Maximum-likelihood phylogenies in Figure 1 and supplementary Supplementary Fig.2, S4 and S5 were reconstructed from the masked alignment using IQ-TREE^78^ v 1.6.12 with GTR model and collapsing near-zero branches (arguments employed: -czb, -m GTR). The phylogenies were rooted in the reference sequence from Wuhan^72^ on 2019-12-24. The phylogenies were annotated and visualized with iTOL^79^ v 4.

The phylogeny in Supplementary Video S1, composed by 10,450 sequences, was build up with Nextstrain pipeline (available at https://github.com/nextstrain/augur) in order to monitor and visualize temporal and geographical transmission of B.1.177.637. This dataset file is available at https://github.com/PathoGenOmics/B.1.177.637_SARS-CoV-2.

### Clusters of transmission involving 1163 and 1167 S amino acid replacements

We used the phylogeny of 10,450 sequences enriched with all sequences mutated in 1163 and 1167 to quantify the minimum number of mutational events involving positions 1163 and 1167 in S protein. We first defined which mutations characterize internal nodes using R packages: tidytree v 0.3.3 and treeio v 1.14.3^80^. We then depicted monophyletic clusters sharing at least one of the two mutations. Transmission clusters were defined as all sequences that: i) are derived from an internal node characterized by the same nucleotide mutation involving 1163 or 1167 amino acid replacements, ii) include more than one sequence, and iii) at least 95% of sequences share the nucleotide mutation. Additionally, redundant nodes were eliminated, keeping the ancestral node of the cluster (harbouring the largest number of leaves). Sequences with at least one mutation but not in clusters were counted as single events of mutation in the phylogeny.

### Structural analysis of 1163 and 1167 S amino acid replacements

The atomic coordinates for S protein in pre-fusion state was retrieved from the CHARMM-GUI COVID-19 Archive (http://www.charmm-gui.org/docs/archive/covid19). The atomic coordinates for S protein in post-fusion sate were retrieved from Protein Data Bank (PDB: 6XRA^18^ and PDB: 6LXT^81^). Mutations were introduced using single mutation tool embedded in COOT^82^and figures were generated with PyMOL (www.pymol.org).

### SARS-CoV-2 pseudotyped vesicular stomatitis virus production, titration, and thermal stability evaluation

Mutations were introduced into a plasmid encoding a codon-optimized S protein^16^ by site directed mutagenesis (see Supplementary table8 for primers). All mutations were verified by Sanger sequencing (see Supplementary table 9 for primers). To evaluate the efficiency of virus production, three transfections in HEK293 cells (CRL-1573 from ATCC) were performed for each plasmid to generate pseudotyped VSV harbouring the indicated S protein^83^. The titers of the virus produced were then assayed by serial dilution, followed by infection of either Vero cells (CCL-81 from ATCC) or A549 cells expressing ACE2 and TMPRSS2 (InvivoGen catalog code a549-hace2tpsa), and counting of GFP positive cells (focus forming units; FFU) at 16 hours post infection. Statistical comparisons were performed by unpaired t-test (R package: stats v 3.6.1) with normalized logarithmic data. For assessing thermal stability, 1000 FFU (as measured on Vero cells) were incubated for 15 minutes at 30.4, 31.4, 33, 35.2, 38.2, 44.8, 47, 48.6 or 49.6°C before addition to Vero cells previously seeded in a 96 well plate (10,000 cells/well). GFP signal in each well was determined 16 hours post-infection using an Incucyte S3 (Essen Biosciences). The mean GFP signal observed in several mock-infected wells was subtracted from all infected wells, followed by standardization of the GFP signal to the mean GFP signal from wells incubated at 30.4°C. Finally, a three parameter log-logistic function was fitted to the data using the drc package v 3.0-1 in R (LL.3 function) and the temperature resulting in 50% inhibition calculated using the drc ED function. Statistical differences in the temperature resulting in 50% reduction of infection was evaluated using the drc EDcomp function. The data and scripts for analysing the temperature resistance of the different mutants is available at https://github.com/PathoGenOmics/B.1.177.637_SARS-CoV-2.

### Evaluation of neutralization by convalescent sera and efficacy of virus particle production

Pseudotyped VSV bearing 20E or B.1.177.637 S variants were evaluated for sensitivity to neutralization by convalescent sera as previously described^83^ with slight modifications. Briefly, 16-hours post-infection, GFP signal in each well was determined using an Incucyte S3 (Essen Biosciences). The mean GFP signal observed in several mock-infected wells was subtracted from all infected wells, followed by standardization of the GFP signal in each well infected with antibody-treated virus to that of the mean GFP signal from wells infected with mock-treated virus. Any negative values resulting from background subtraction were arbitrarily assigned a low, non-zero value (10^-5^). The serum dilutions were then converted to their reciprocal, their logarithm (Log10) was taken, and the dose resulting in 50% (ID50) or 80% (ID80) reduction in GFP signal was calculated in R using the drc package v 3.0-1. A two-parameter log-logistic regression (LL2 function) was used for all samples except when a three-parameter logistic regression provided a significant improvement to fit, as judged by the ANOVA function in the drc package (e.g. p < 0.05 following multiple testing correction using the Bonferroni method). The script for calculating the ID50 and ID80 as well as the standardized GFP signal for each condition is available at https://github.com/PathoGenOmics/B.1.177.637_SARS-CoV-2. For the first wave, serum samples and data from patients included in this study were provided by the Consorcio Hospital General de Valencia Biobank, integrated in the Valencian Biobanking Network, and they were processed following standard operating procedures with the appropriate approval of the Ethics and Scientific Committees. All first wave samples were obtained from donors that were admitted to the intensive care unit and were collected during April 2020. For the second wave donors, sera were obtained (October 2020) from severe COVID-19 patients requiring inpatient treatment at Hospital Universitario y Politécnico La Fe de Valencia. Similarly, samples from immunized donors were collected at Hospital Universitario y Politécnico La Fe de Valencia from hospital health-workers, with no previous history of SARS-COV-2 infection, and after receiving a second dose of Pfizer-BioNTech COVID-19 vaccine (BNT162b2; February 2021). All samples from Hospital Universitario y Politécnico La Fe de Valencia were collected after informed written consent and the project has been approved by the ethical committee and institutional review board (registration number 2020-123-1).

## Data Availability

Data referred to this manuscript is available as supplemental material, and code can be found at https://github.com/PathoGenOmics/B.1.177.637_SARS-CoV-2. The analysis pipeline used to map and analyze the sequences is available at https://gitlab.com/fisabio-ngs/sars-cov2-mapping

https://gitlab.com/fisabio-ngs/sars-cov2-mapping

## Acknowledgments

We want to particularly acknowledge the patients and the Consorcio Hospital General de Valencia Biobank integrated in the Valencian Biobanking Network for their collaboration, as well as the patients and hospital staff at Hospital Universitario y Politécnico La Fe de Valencia. In addition, the authors would like to thank Gert Zimmer (Institute of Virology and Immunology, Mittelhäusern/Switzerland), Stefan Pohlmann, and Markus Hoffmann (German Primate Center, Infection Biology Unit, Goettingen/Germany) for providing the reagents required for the generation of VSV pseudotyped viruses and the codon-optimized S plasmid. Also, we want to acknowledge all the efforts from different laboratories and authorities submitting all possible sequences of SARS-CoV-2 worldwide and making them available on the GISAID platform.

## Author contributions

PRR: Conceptualization, Methodology, Formal Analysis, Investigation, Visualization, Writing Original Draft.

CFG: Conceptualization, Methodology, Formal Analysis, Writing Original Draft.

ACO: Formal Analysis, Review and edit draft.

MGL: Formal Analysis, Review and edit draft.

SJS: Software, Validation, Review and edit draft.

ICM: Methodology, Resources, Review and edit draft.

PRH: Investigation, Review and edit draft.

MTP: Methodology, Resources, Review and edit draft.

MAB: Investigation, Methodology, Review and edit draft.

GD: Methodology, Software, Data Curation, Review and edit draft.

LLM: Methodology, Project Administration, Review and edit draft.

MG: Resources, Review and edit draft.

MMA: Resources, Review and edit draft.

MDG: Resources, Review and edit draft.

JLP: Resources, Review and edit draft.

FG: Funding, Project Administration, Supervision, Review and edit draft.

IC: Funding, Project Administration, Supervision, Review and edit draft.

AM: Formal analysis, Writing Original Draft.

RG: Conceptualization, Methodology, Formal Analysis, Writing Original Draft, Supervision, Funding.

MC: Conceptualization, Methodology, Investigation, Formal Analysis, Writing Original Draft, Supervision, Funding.

## List of SeqCOVID consortium members

Iñaki Comas, Fernando González-Candelas, Galo A. Goig-Serrano, Álvaro Chiner-Oms, Irving Cancino-Muñoz, Mariana Gabriela López, Manoli Torres-Puente, Inmaculada Gómez, Santiago Jiménez-Serrano, Lidia Ruiz-Roldán, María Alma Bracho, Neris García-González, Llúcia Martínez Priego, Inmaculada Galán-Vendrell, Paula Ruiz-Hueso, Griselda De Marco, Mª Loreto Ferrús Abad, Sandra Carbó-Ramírez, Mireia Coscollá, Paula Ruiz Rodríguez, Giuseppe D’Auria, Francisco Javier Roig Sena, Hermelinda Vanaclocha Luna, Isabel San Martín Bastida, Daniel García Souto, Ana Pequeño Valtierra, Jose M. C. Tubio, Fco. Javier Temes Rodríguez, Jorge Rodríguez-Castro, Martín Santamarina García, Nuria Rabella Garcia, Ferrán Navarro Risueño, Elisenda Miró Cardona, Manuel Rodríguez-Iglesias, Fátima Galán-Sanchez, Salud Rodríguez-Pallares, María de Toro, María Pilar Bea-Escudero, José Manuel Azcona-Gutiérrez, Miriam Blasco-Alberdi, Alfredo Mayor, Alberto L. GarcÍa-Basteiro, Gemma Moncunill, Carlota Dobaño, Pau Cisteró, Oriol Mitjà, Camila González-Beiras, Martí Vall-Mayans, Marc Corbacho-Monné, Andrea Alemany, Darío García de Viedma, Laura Pérez-Lago, Marta Herranz, Jon Sicilia, Pilar Catalán, Julia Suárez, Patricia Muñoz, Cristina Muñoz-Cuevas, Guadalupe Rodríguez Rodríguez, Juan Alberola Enguídanos, Jose Miguel Nogueira Coito, Juan José Camarena Miñana, Antonio Rezusta López, Alexander Tristancho Baró, Ana Milagro Beamonte, Nieves Martínez Cameo, Yolanda Gracia Grataloup, Elisa Martró, Antoni E. Bordoy, Anna Not, Adrián Antuori, Anabel Fernández, Nona Romaní, Rafael Benito Ruesca, Sonia Algarate Cajo, Jessica Bueno Sancho, Jose Luis del Pozo, Jose Antonio Boga Riveiro, Cristián Castelló Abietar, Susana Rojo Alba, Marta Elena Álvarez Argüelles, Santiago Melón García, Maitane Aranzamendi Zaldumbide, Óscar Martínez Expósito (), Mikel Gallego Rodrigo (), Maialen Larrea Ayo (), Nerea Antona Urieta (), Andrea Vergara Gómez, Miguel J Martínez Yoldi, Jordi Vila Estapé, Elisa Rubio García, Aida Peiró-Mestres, Jessica Navero-Castillejos, David Posada, Diana Valverde, Nuria Estévez, Iria Fernández-Silva, Loretta de Chiara, Pilar Gallego, Nair Varela, Rosario Moreno Muñoz, Mª Dolores Tirado Balaguer, Ulises Gómez-Pinedo, Mónica Gozalo Margüello, Mª Eliecer Cano García, José Manuel Méndez Legaza, Jesús Rodríguez Lozano, María Siller Ruiz, Daniel Pablo Marcos, Antonio Oliver, Jordi Reina, Carla López-Causapé, Andrés Canut Blasco, Silvia Hernáez Crespo, Mª Luz Cordón Rodríguez, Mª Concepción Lecaroz Agara, Carmen Gómez González, Amaia Aguirre Quiñonero, José Israel López Mirones, Marina Fernández Torres, Mª Rosario Almela Ferrer, José Antonio Lepe Jiménez, Verónica González Galán, Ángel Rodríguez Villodres, Nieves Gonzalo Jiménez, Mª Montserrat Ruiz García, Antonio Galiana Cabrera, Judith Sánchez-Almendro, Gustavo Cilla Eguiluz, Milagrosa Montes Ros, Luis Piñeiro Vázquez, Ane Sorrarain, José María Marimón Ortiz de Zarate, Mª Dolores Gómez Ruiz, Eva González Barberá, José Luis López Hontangas, José María Navarro-Marí, Irene Pedrosa Corral, Sara Sanbonmatsu Gámez, M. Carmen Perez Gonzalez, Francisco Javier Chamizo López, Ana Bordes Benítez, David Navarro Ortega, Eliseo Albert Vicent, Ignacio Torres, Mª Isabel Gascón Ros, Cristina Torregrosa Hetland, Eva Pastor Boix, Paloma Cascales Ramos, Begoña Fuster Escrivá, Concepción Gimeno Cardona, María Dolores Ocete Mochón, Rafael Medina González, Julia González Cantó, Olalla Martínez Macias, Begoña Palop Borrás, Inmaculada de Toro Peinado, Mª Concepción Mediavilla Gradolph, Mercedes Pérez Ruiz, Oscar González-Recio, Mónica Gutiérrez-Rivas, Encarnación Simarro Córdoba, Julia Lozano Serra, Lorena Robles Fonseca, Adolfo de Salazar, Laura Viñuela, Natalia Chueca, Federico García, Cristina Gomez-Camarasa, Ana Carvajal, Vicente Martín, Juan Fregeneda, Antonio J. Molina, Héctor Arguello, Tania Fernandez-Villa, Amparo Farga Martí, Rocío Falcón, Victoria Domínguez Márquez, José Javier Costa Alcalde, Rocío Trastoy Pena, Gema Barbeito Castiñeiras, Amparo Coira Nieto, María Luisa Pérez del Molino Bernal, Antonio Aguilera, Anna M. Planas, Álex Soriano, Israel Fernández-Cádenas, Jordi Pérez-Tur, Mª Ángeles Marcos Maeso, Carmen Ezpeleta Baquedano, Ana Navascués Ortega, Ana Miqueleiz Zapatero, Manuel Segovia Hernández, Antonio Moreno Docón, Esther Viedma Moreno, Jesús Mingorance, Juan Carlos Galán Montemayor, Iván Sanz Muñoz, Diana Pérez San José, Maria Gil Fortuño, Juan B. Bellido Blasco, Alberto Yagüe Muñoz, Noelia Henández Pérez, Helena Buj Jordá, Óscar Pérez Olaso, Alejandro González Praetorius, Aida Esperanza Ramírez Marinero, Eduardo Padilla León, Alba Vilas Basil, Mireia Canal Aranda, Albert Bernet Sánchez, Alba Bellés Bellés, Eric López González, Iván Prats Sánchez, Mercè García González, Miguel Martínez Lirola, Maripaz Ventero Martín, Carmen Molina Pardines, Nieves Orta Mira, María Navarro Cots, Inmaculada Vidal Catalá, Isabel García Nava, Soledad Illescas Fernández-Bermejo, José Martínez-Alarcón, Marta Torres-Narbona, Cristina Colmenarejo, Lidia García-Agudo, Jorge Alfredo Pérez García, Martín Yago López, María Ángeles Goberna Bravo, Carolina Pla Cortes, Noelia Lozano Rodríguez, Nieves Aparici Valero, Sandra Moreno Marro, Agustín Irazo Tatay, Isabel Mariscal Pieper, Mª Pilar Ramos, Mónica Parra Grande, Bárbara Gómez Alonso, Francisco José Arjona Zaragozí, Amparo Broseta Tamarit, Juan José Badiola Díez, Alicia Otero García, Eloísa Sevilla Romeo, Belén Marín González, Mirta García Martínez, Marina Betancor Caro, Diego Sola Fraca, Sonia Pérez Lázaro, Eva Monleón Moscardó, Marta Monzón Garcés, Cristina Acín Tresaco, Rosa Bolea Bailo, Bernardino Moreno Burgos, Amparo Broseta Tamarit, Carlos Gulin Blanco

## Supplementary Material

**Supplementary video 1.** Geographical transmission of B.1.177.637 visualized with Nextstrain build.

## Supplementary Tables

**Supplementary table 1.** Accession numbers for analysed sarbecoviruses.

**Supplementary table 2.** Defining SNPs for B.1.177.637, B.1.177.637.V2, and VOI1163.654.

**Supplementary table 3.** Accession numbers of sequences of interest for mutated amino acids 484, 501, 1163 and 1167 of the S protein.

**Supplementary table 4.** Accession numbers of 5,017 Spanish sequences from SeqCOVID-SPAIN consortium.

**Supplementary table 5.** Accession numbers of 270,869 analysed sequences.

**Supplementary table 6.** Accession numbers of 10,450 analysed sequences.

**Supplementary table 7.** Accession numbers of 3,067 analysed sequences belonging to 20I/501Y.V1.

**Supplementary table 8.** Primers for site directed mutagenesis of plasmid encoding codon-optimized S protein.

**Supplementary table 9.** Primers for sanger sequencing to detect mutations of interest.

## Supplementary Figures

**Supplementary Figure 1.**
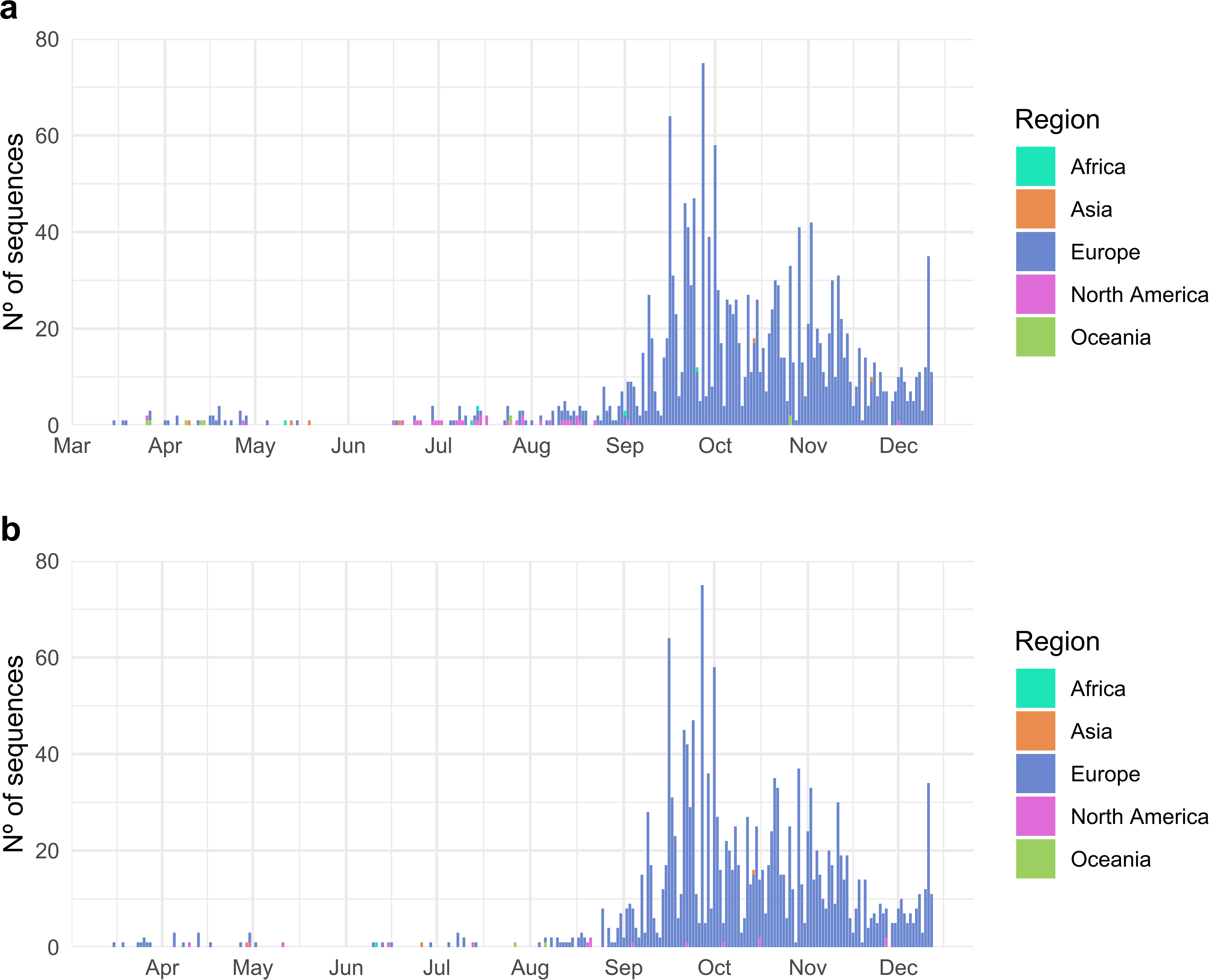
Temporal distribution of mutated samples coloured by region. **a.** Distribution of S protein amino acid replacement D1163Y over the pandemic (N=1874). **b.** Distribution of S protein amino acid replacement G1167V over time (N=1708).

**Supplementary Figure 2.**
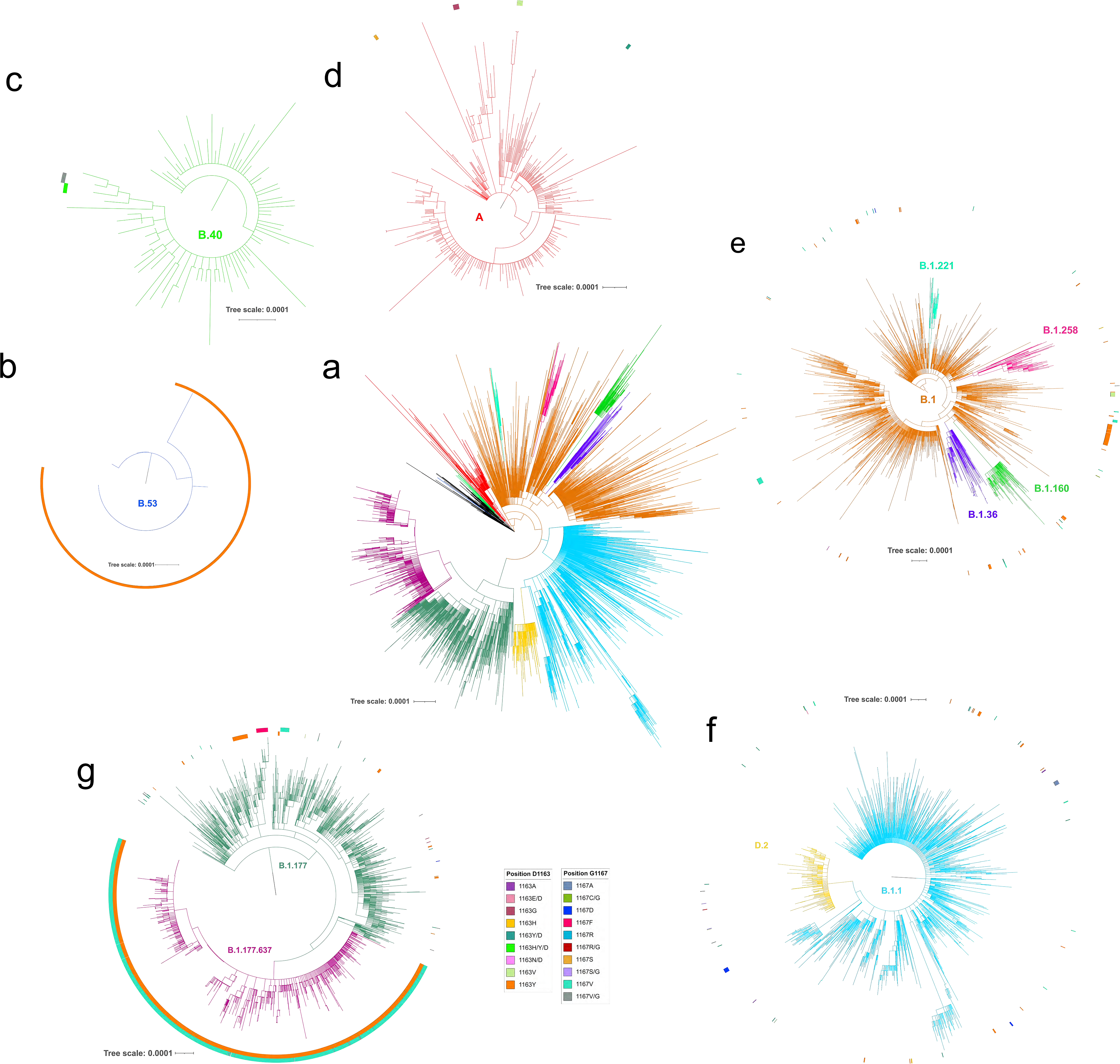
Maximum-likelihood phylogenies for different PANGO lineages. **a.** Complete phylogeny coloured by the PANGO lineages. **b.** Phylogeny of B.53 lineage. The circle represents sequences with D1163 amino acid replacement. **c.** Phylogeny of B.53 lineage. The inner circle represents sequences with D1163 amino acid replacements, and the external circle represents sequences with G1167 amino acid replacements. **d.** Phylogeny of A lineage. The circle represents sequences with D1163 amino acid replacements. **e.** Phylogeny of B.1 and derived lineages. The inner circle represents sequences with D1163 amino acid replacements, and the external circle represents sequences with G1167 amino acid replacements. **f.** Phylogeny of B.1.1 and derivative D.1 lineages. The inner circle represents sequences with D1163 amino acid replacements, and the external circle represents sequences with G1167 amino acid replacements. **g.** Phylogeny of B.1.177 and B.1.177.637 lineages. The inner circle represents sequences with D1163 amino acid replacements, and the external circle represents sequences with G1167 amino acid replacements. The scale bar of each indicates the number of nucleotide substitutions per site. The legend of positions 1163 and 1167 is common for all represented panels.

**Supplementary Figure 3.**
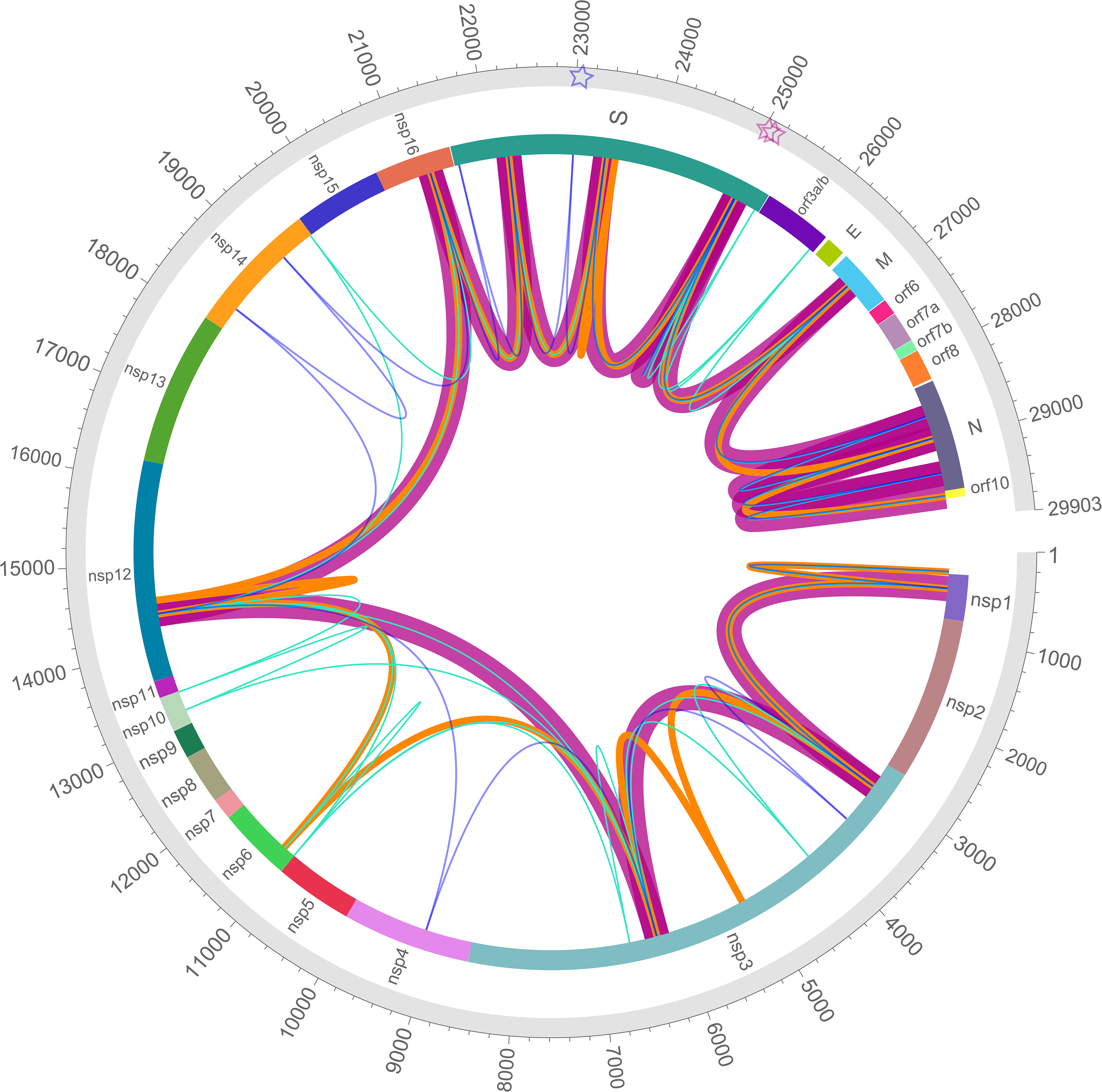
Whole genome mutated positions in genotypes with changes 1163Y and 1167V of the S protein. B.1.177.637 is coloured in magenta and cluster 1163.654 is coloured in orange. Other less frequent genotypes (found in at least 20 sequences) that include changes in S position 1163 and/or 1167 are coloured in turquoise. Cluster 1163.654 is coloured in navy blue. Line width is proportional to the frequency of the genotype. Sites 1163 and 1167 in the S protein are indicated by magenta star symbols, and position 484 in S, whose mutations are associated with antigenicity changes, is indicated by a navy blue star.

**Supplementary Figure 4.**
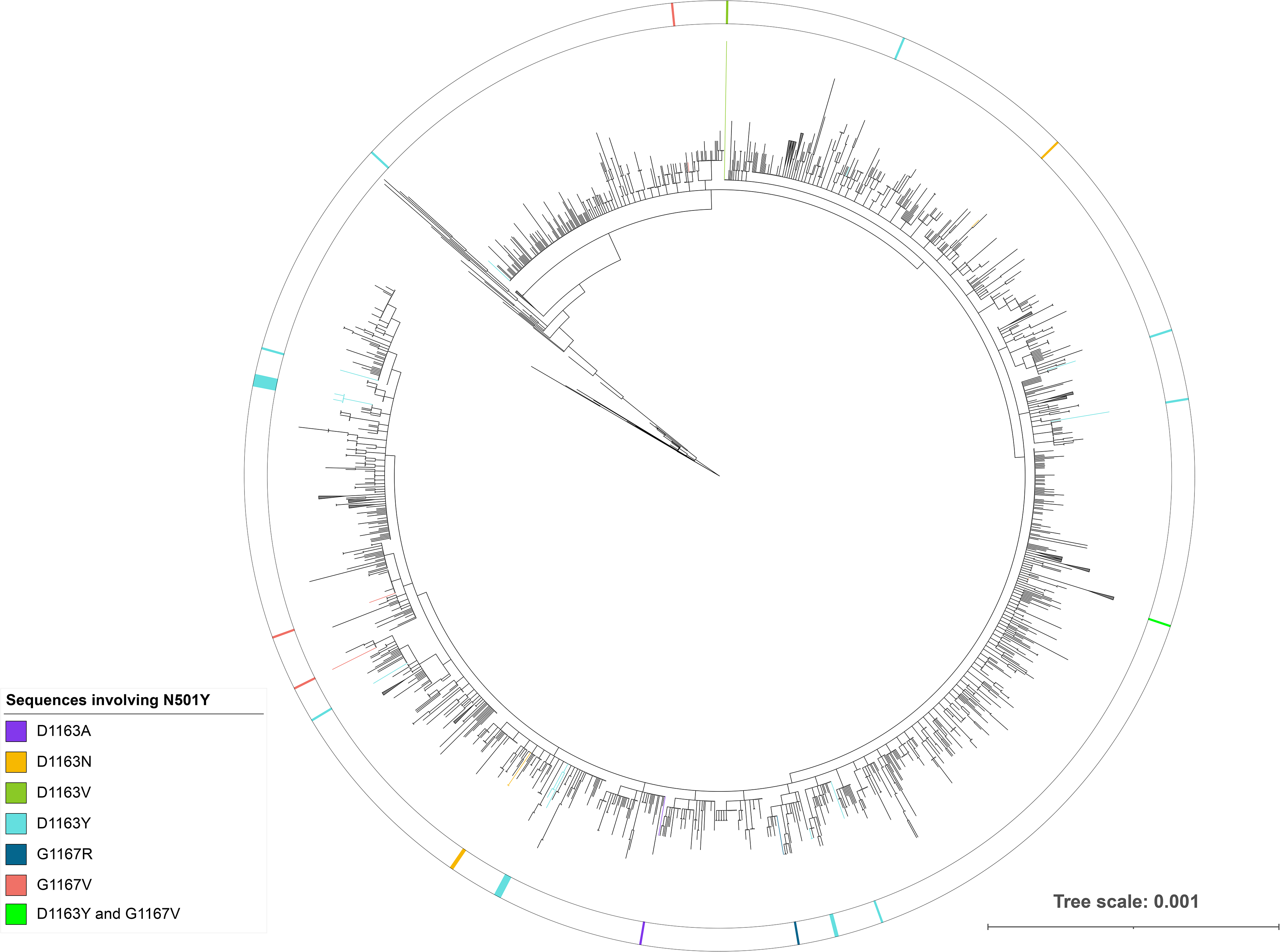
Maximum-likelihood phylogeny of 3,067 genomes belonging to 20I/501Y.V1, rooted with Wuhan reference sequence. Mutated sequences with 1163 and/or 1167 amino acids of the S protein are coloured in the circle. The scale bar indicates the number of nucleotide substitutions per site. The biggest clades are collapsed with isosceles triangles.

**Supplementary Figure 5.**
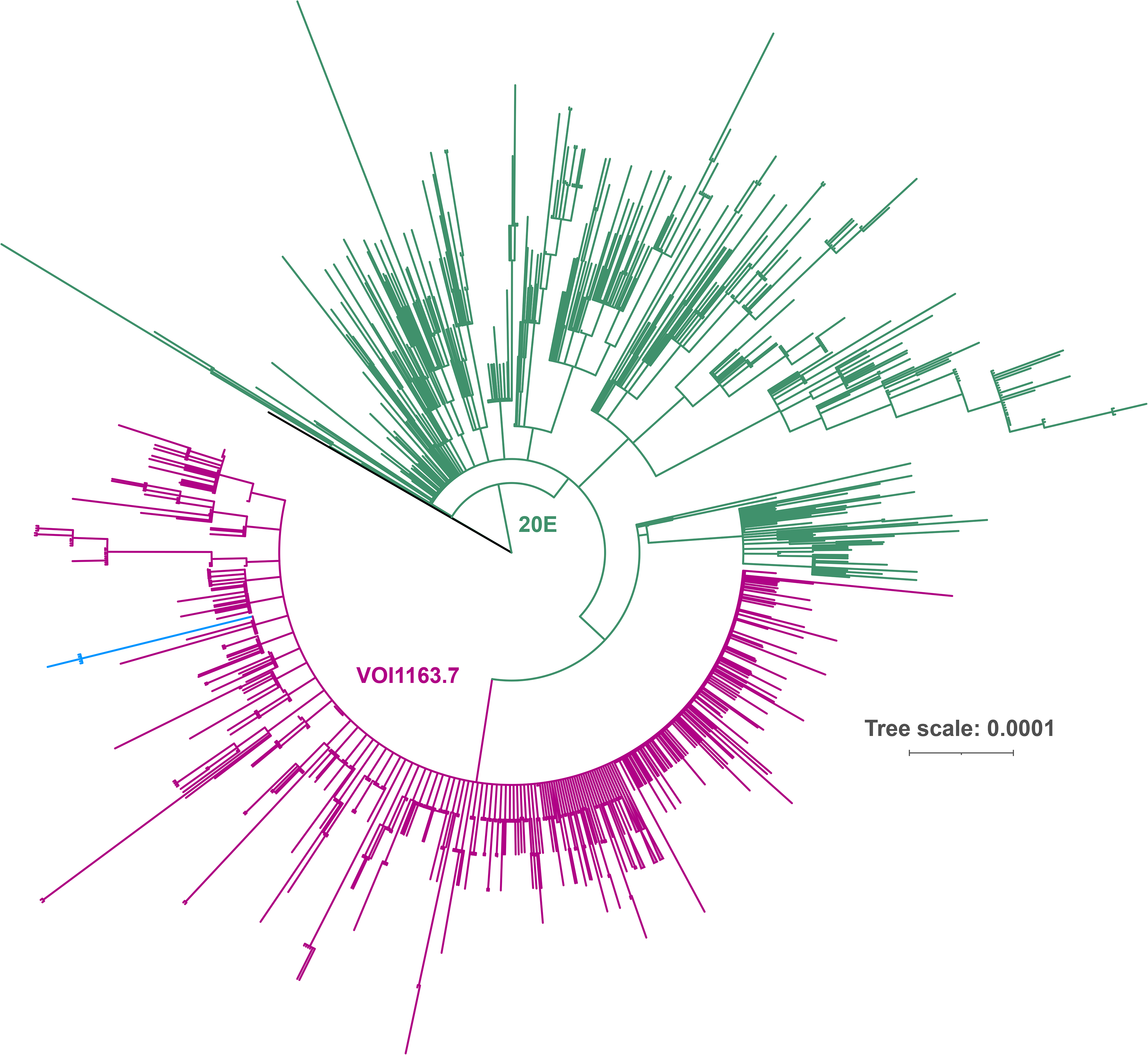
Maximum-likelihood phylogeny of 3,266 SARS-CoV-2 genomes representing 20E clade rooted with Wuhan reference sequence. Sequences from B.1.177.637 are coloured in magenta, sequences not identified as B.1.177.637 are coloured in green, and cluster B.1.177.637.V2 (S protein amino acid replacements: A222V, D614G, E484K, D1163Y, and 141-144Del) is coloured in blue. The scale bar indicates the number of nucleotide substitutions per site. The biggest clades are collapsed, represented as isosceles triangles.

**Supplementary Figure 6.**
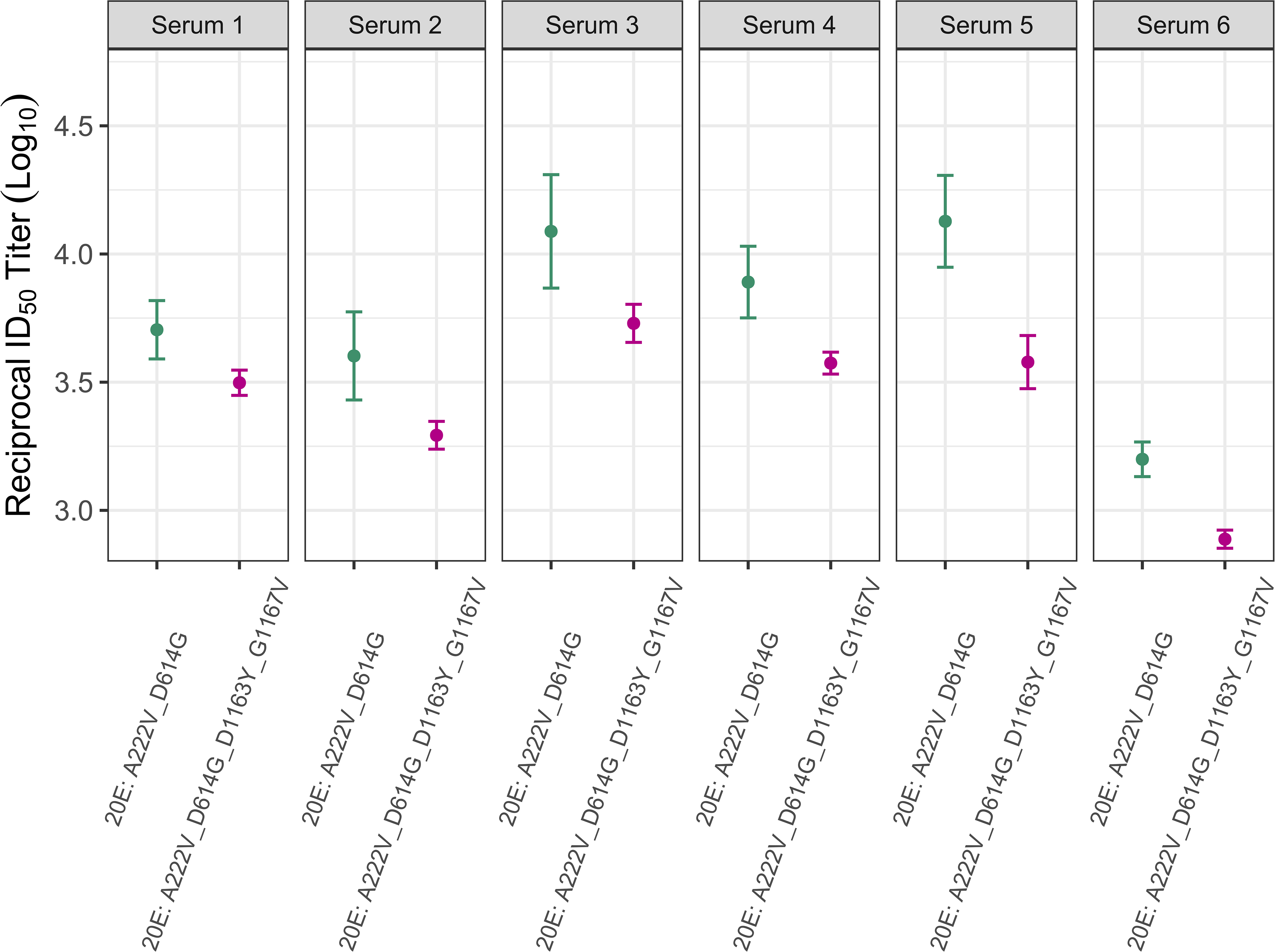
Neutralization of the different mutated S protein variants by convalescent sera from six individuals infected during the first epidemic wave. The reciprocal titer at which each of the different convalescent sera neutralizes the different variants by 50% is indicated. Plotted are the mean and standard error of (n=3).

